# Engaging Young People with Experience of Mental Health Needs: Reflections from Young People and Outputs from a Resource Development Project

**DOI:** 10.1101/2025.03.06.25323541

**Authors:** Zoë Haime, Charlotte Carney, Myles Jay-Linton, Helen Bould, Lucy Biddle

## Abstract

Recommendations advise mental health practitioners to discuss problematic online use with children and young people. However, barriers such as knowledge gaps and low confidence in initiating discussions often prevent these conversations from happening. The Digital Dialogues project used a knowledge exchange approach co-creating resources with young people to support professionals in overcoming these challenges. This paper details the project design and reflects on the perspectives of the young people involved.

The project was guided by the ‘children and young people have ownership’ model of co-creation. 11 participants were purposively sampled to take part in the Digital Dialogues Young Persons Group (DDYPG) and were actively involved in the study workshops, creative tasks, and resource design and development. Six DDYPG members took part in interviews, and two also completed an anonymous survey, evaluating their time in the DDYPG. Thematic analysis was used to explore data from interviews and qualitative survey responses.

The DDYPG successfully created several resources to support practitioners in addressing problematic online use with young people. Reflections from DDYPG members showed that creative engagement, meaningful involvement, and peer interactions were key motivators for participation and led to benefits including feelings of empowerment and personal development. Anxiety, time demands, and potential exposure to triggering content could act as barriers. However, structured tasks, positive rapport with researchers, and flexible participation helped to mitigate these challenges.

Findings highlight ethical considerations and potential strategies for involving young people in resource development research projects in the future.

## Introduction

Online use can bring positive opportunities to children and young people (CYP), including learning, connectedness, and fun. In relation to their to their mental health, it can also encourage access to helpful information and peer support [1–2]. However, there are concerns around risks associated with online use in CYP. For instance, links exist between engaging with harmful or distressing images and maladaptive behaviours including self-harm and disordered eating [3–4]. Additionally, negative online experiences have been significantly associated with increased psychiatric symptoms in CYP [5].

As a result, recommendations have been made for mental health professionals (MHPs) working with CYP to support their online use. This includes advice from RCPsych [6] for psychiatrists to enquire about online use during all consultations with young people. Research has also shown a willingness amongst MHPs to discuss this topic with young people [7], but several barriers including knowledge gaps, time constraints, and a lack of confidence, prevent them from doing so [8–9]. As a result, guidelines have been developed, such as the Good Practice Indicators (GPIs) which act as advice for those managing these conversations in mental health practice [10]. However, little is known about their implementation in practice, and MHPs lack practical resources to navigate these conversations effectively.

The Digital Dialogues project aimed to use a knowledge exchange approach [11] to develop additional resources for MHPs, aiding their discussions with young people around online use. Firstly, in an evidence-synthesis phase, we conducted two nationwide surveys to enquire about 1) what resources and training MHPs want and need [12], and 2) what thoughts and feelings CYP have about professionals working with them around this topic. Secondly, in a resource development phase, we collaborated with young people, using creative methods such as art, poetry, and drama to engage them and allow for self-expression of thoughts and ideas through a variety of means [13]. During this phase, we established the Digital Dialogues Young Person’s Group (DDYPG), providing a space for young people with lived and living experience of mental health needs to contribute to Digital Dialogues in member roles.

This paper aims to outline and evaluate ways DDYPG members were involved as members in the Digital Dialogues resource development phase. We present details of the workshops, creative tasks, and project processes, to demonstrate how we involved and engaged young people, alongside interview data in which participants reflect on their experiences.

## Method

### Collaborative Approach

We aimed to collaborate with CYP with lived and living experiences of mental health needs, to generate ideas for potential resources based on their experiences and perspectives. Drawing on the *Guidelines for Research with Children and Young People* [14] we focused on approaching the study with the ‘CYP have ownership’ model of involvement. By doing so, we hoped to provide CYP with agency over the research process and to embed them as research team members, whilst providing guidance, support, and navigation from the trained research team [15].

### Digital Dialogues Young Person’s Group Recruitment

A digital recruitment advert was distributed via various young people’s groups, including Arts Emergency, Partnership for Young London, and the National Youth Agency, as well as specific mental health organisations including McPin, OCD Youth, BDD Foundation, What Works Wellbeing, Mental Movement Magazine, and Beyond. Additionally, the advert was shared through the Epigram University of Bristol Student Newspaper, and relevant societies at universities across the UK, including the ThinkMental Kings College London Society, Beat This Together University of Bristol Society, and StudentMinds University of York Society.

Potential DDYPG members completed an expression of interest form, detailing their name, email address, age, and creative interests. They were then assessed against eligibility criteria for involvement:

- 14-25yrs old
- lived or living experience of engaging online regarding their own mental health
- willingness to participate for up to 7 months
- access to a stable internet connection
- adequate understanding of the English language
- currently residing in the UK

Study information sheets were sent to all (N=45) eligible potential participants, and of those who continued to express an interest, 11 were purposively sampled and invited for an individual introductory session with researcher ZH. Purposive sampling was employed to ensure a diverse population, selecting DDYPG members with varying demographics as well as creative interests, to enrich the range of perspectives within the study. During introductory sessions potential members were able to ask questions, learn about the safety plan and consent process, and provide brief information regarding their online use and mental health experiences. Following these sessions, all 11 potential DDYPG members consented to take part in the DDYPG. During this process, members gave consent for their contributions to be used and shared in resources, and provided separate consent for any potential sharing of their creative work. At this point, they also completed a survey that informed researchers of specific triggers they may have related to mental health content. Recruitment took place between October and November 2023, and was closed once all members had provided consent.

### Ethical Considerations

Ethical approval was given by the Faculty of Health Research Ethics Committee at the University of Bristol (reference number: 15930). Although this was public engagement work, ethical approval was sought due to the involvement of vulnerable young people with mental health needs, the planned creative outputs, and our intention to evaluate the collaborative work. We wanted to ensure group members were appropriately safeguarded, and fully informed around their rights regarding the creation and sharing of materials during the study.

As part of our ethical approach, we required DDYPG members to complete an individual safety plan (Supplementary Material – Appendix A). In this plan, members provided details of an emergency contact and their GP, to be used if researchers identified an immediate risk of harm to themselves or others. Additionally, they could create a personalised care plan and access a range of wellbeing resources. Researchers also followed a distress protocol during the project, including following up with members individually after each workshop.

### Online Platform Communication

As part of this study we set up a private server on the online communication platform, Discord. The Discord platform supports discussions as well as having features enabling file-sharing. Additionally, Discord is a popular platform amongst young people that has been shown to enhance digital collaboration [16]. We believed this would be an effective way to encourage conversations amongst young people, engagement with study materials, and sharing of information. All DDYPG members and Digital Dialogues researchers were invited to join if they wished, with ZH moderating content. Platform discussions were restricted during non-working hours.

### Digital Dialogues Young Person’s Group Procedure

Three DDYPG workshops (Table 1) took place via online videoconferencing platform Teams, between November 2023 and January 2024. All workshops were audio-recorded and audio was transcribed by ZH, who then created and shared a workshop summary with all DDYPG members.

**Table 1.** Task Aims and Instructions.

| Task | Aim | Instructions |
| --- | --- | --- |
| **Task 1: GPI Creative Work** | Create a visual or written piece reflecting on a Good Practice Indicator [10]. | Members received a welcome pack with creative materials and two suggested methods: (1) **Erasure poems** (using pages from books)<br>(2) **Smartphone template drawings**.<br>They could also use their own artistic style. Creations were shared in Workshop 1. |
| **Task 2: Survey** | Gather young people's views on online culture, mental health, and digital communication. | A short survey exploring emoji meanings, mental health platforms, online trends, influencers, and experiences with MHPs. Results were discussed in Workshop 2. |
| **Task 3: Search History Poems** | Use list-style poetry to reflect on online searches and mental health experiences. | Inspired by poems by Vuong [43] and Boensch [44], members created poems using real or fictional search histories to narrate their online journeys. Results were discussed in Workshop 3. |
| **Task 4: Character Development** | Develop a character for a mental health video resource. | During Workshop 3, members brainstormed a ‘day in the life’ concept. They completed character profiles covering backstory, social media habits, daily experiences, and an MHP interaction (both positive and negative scenarios). |

Where young people were unable to attend, or preferred not to be involved in workshops, they were given the opportunity to take part in alternative ways, such as involvement in discussions over Discord, or creating, revising and editing documents and resources.

Throughout the project young people also took part in several creative tasks (Table 2). Instructions for tasks were shared via Discord and email, and for Task 1 they were posted to a given address, alongside some creative materials. Creative work was used to encourage young people’s involvement in discussions related to their experiences, and to encourage idea generation for the resulting resources [17].

**Table 2.** Details of resources developed during Digital Dialogues.

| Resource | Description | DDYPG Involvement |
| --- | --- | --- |
| Flash Cards & Road Maps | 20 flash cards representing a safety mechanism related to online use with a lived experience account detailing CYP experience of using it. | 3 members contributed lived experience content.<br>5 selected the flash card design. |
|  | Road maps outlined a potential online scenario, and the relevant flash cards for MHPs to use with it. | • 1 conceptualised the creation of roadmaps and designed them.<br>• 1 reviewed and edited the roadmaps. They also reviewed researcher changes and made amendments. |
| **Content Creator Advice Poster & Additional Information** | Poster outlining considerations for MHPs when CYP are following a content creator who posts mental health-related content. A document with more detailed guidance was also created. | • 1 member conceptualised and designed the poster. They also reviewed researcher changes and made amendments.<br>• 1 developed a detailed information document to complement the poster. |
| **Comfortable Conversations with MHPs Poster & Question Prompt Bank** | Poster outlining strategies MHPs could use to help CYP feel comfortable during conversations around their online use.<br>The question prompt bank offers MHPs a curated list of important questions to ask CYP. | • 8 members conducted content analysis of a CYP survey.<br>• 1 member conceptualised and designed the poster. They also reviewed researcher changes and made amendments.<br>• 1 developed the question prompt bank. They also reviewed researcher changes and made amendments. |
| **Video** | Video portraying a "day in the life" of a YP whose behaviours and mood are both positively and negatively impacted by her online engagement, particularly in relation to eating. | • 8 members produced character descriptions (Task 4), which researchers merged to form the lead character.<br>• 2 reviewed, discussed, and edited the script (made by the research team).<br>• 1 member helped audition potential actors.<br>• 8 provided voiceovers for the video. |
| **Virtual Exhibition** | The virtual exhibition serves as a platform to display the creative outputs produced by DDYPG members throughout the study. | • 7 members reviewed the virtual exhibition and gave feedback for changes. |

After the final workshop, in which shared decision-making allowed researchers and DDYPG members to outline what resources the group would create, ZH contacted DDYPG members individually about their involvement (Table 3). In some cases, members also approached ZH with ideas for resources to develop. Members worked on resources independently or in groups, alongside input from researchers where indicated as necessary by the young people, between Jan 2024-May 2024.

Seven DDYPG members also received training in content analysis methods and contributed to a separate manuscript, and three made content for Digital Dialogues presentations at conferences. Additionally, creative outputs by DDYPG members were displayed in a virtual exhibition that members reviewed and provided feedback on (https://www.artsteps.com/view/65d476116d3d265cfa4445f2)

Members were reimbursed for their involvement in the study with payments of £25 per hour, as vouchers for an online shop.

Following the creation of the resources, Digital Dialogues II has been funded, and commenced in November 2024. This project aims to develop a training package and session for MHPs, that incorporates the Digital Dialogues resources. Dissemination of the Digital Dialogues resources is therefore ongoing, with DDYPG members being consulted on an ongoing basis.

### Workshops

DDYPG members took part in three online workshops designed to create a space for young people to share their experiences with online use and mental health, whilst also considering the perspectives of MHPs. Prior to each workshop, DDYPG members received details about pre-workshop tasks, what would happen during the workshop, and post-workshop follow-ups (Supplementary Material - Table A). The primary aim of the workshops was to work towards idea generation for resource creation and prepare members to bring their own experiences and insights into the resource development phase.

### Tasks

Tasks were completed to help young people reflect on their personal experiences, with creative methods used to allow novel ways of self-expression. Ultimately, the information gained through workshop discussions of tasks, informed the conception and development of resources, ensuring that the perspectives and experiences of all DDYPG members were incorporated. Tasks are described in Table 1, and in detail in Supplementary Material – Table B.

### Study Flow

The participant study flow is detailed in Figure 1.

**Figure 1 –.**
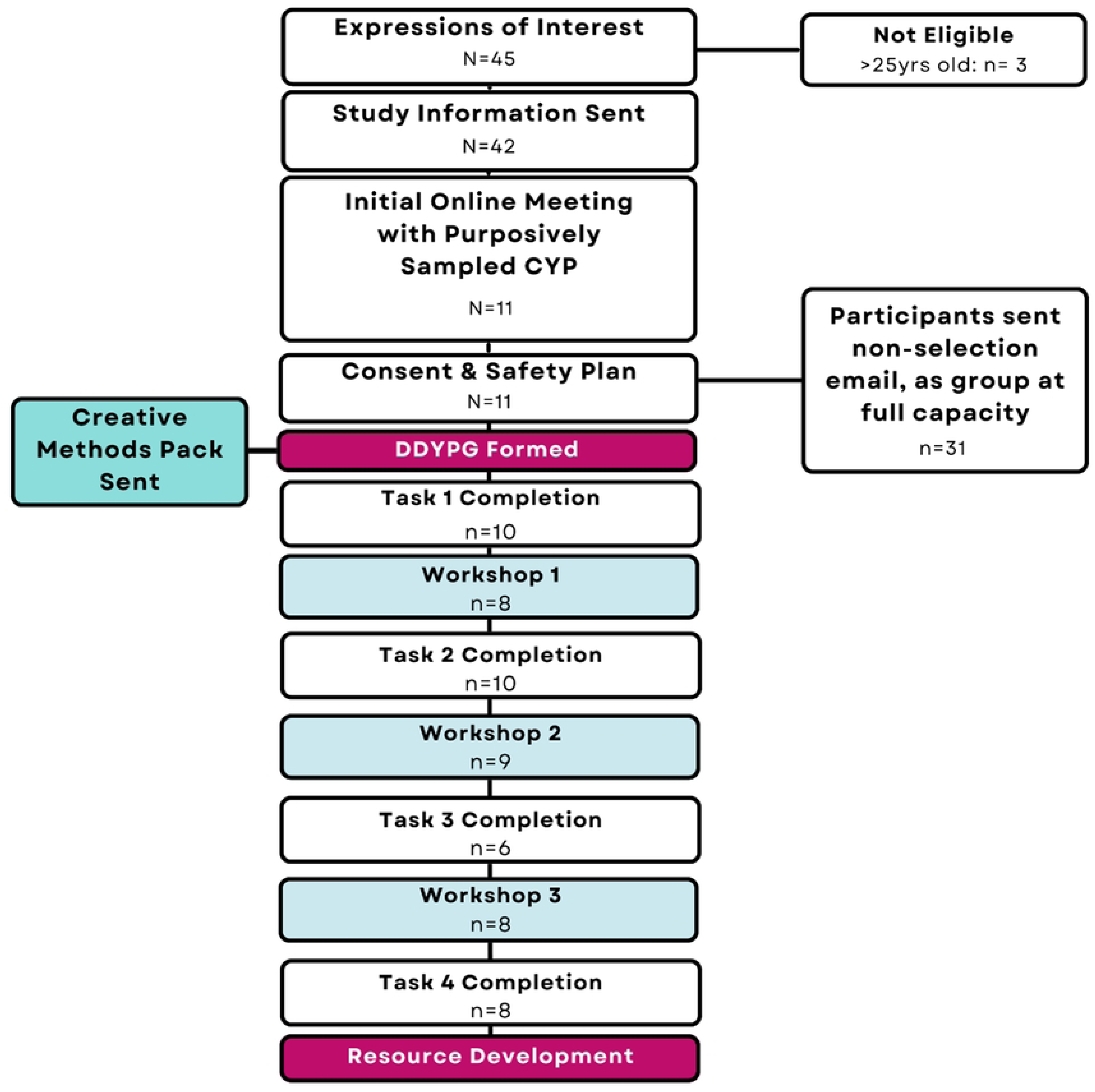
Digital Dialogues Young Person Group Study Flow. **Notes:** CYP = Children and Young People

### Evaluation of Involvement in Digital Dialogues

All DDYPG members (N=11) were invited to evaluate their time in the study via a one-to-one online interview and by completing an anonymous survey with similar questions, allowing for additional feedback. Interviews were semi-structured and conducted by ZH, using a topic guide exploring the positives and negatives of involvement in the DDYPG, suggested changes for the DDYPG, reflections on specific workshops and tasks, and opinions on the resulting resources. Additionally, all members were invited to complete an anonymous survey. Audio from interviews was transcribed by ZH.

Researcher CC joined the Digital Dialogues project after the evaluation interviews had been conducted and carried out the initial coding of transcripts using thematic analysis [18]. The involvement of CC ensured a layer of analysis from a researcher not involved with the data collection, helping to enhance rigor and reduce bias. Thematic analysis was chosen for its flexibility and systematic approach. After initial coding, CC then organised the codes and generated themes, which went through an iterative process following feedback from ZH. There was then a member checking phase, where three DDYPG members were invited to review the resulting data to ensure accurate representation, this led to minor refinements such as adding detail to positive changes in online behaviour, and including more information on flexibility towards member involvement.

## Results

### DDYPG Member Demographics

Eight Members identified as female, two as male, and one as non-binary. All were aged between 18-24yrs and based in the UK. Members had experience with a range of mental health difficulties including anxiety, depression, personality disorder, eating disorders, and OCD.

### Resources

Table 2 shows details of the resources that were conceptualised, designed, and created during this project, with contributions by the DDYPG members detailed. Resources were created simultaneously, over a period of five months to ensure young people had time to contribute alongside other commitments. Researchers provided feedback and edits on resources, which were then reviewed and amended by the group members before a resource was finalised. Finalised resources can be viewed at www.digitaldialogues.co.uk.

### Evaluation

Six DDYPG members completed evaluation interviews and 2 took part in the anonymous survey. Results from the thematic analysis of the qualitative response data are presented below by theme.

#### Reflecting on Involvement in Creative Tasks

DDYPG members gave mixed feedback when reflecting on Task 1 (Table 2). When approaching the task, one (P03) felt completely unable to complete it, and others expressed struggling, feeling they lacked the necessary artistic ability:

> *‘I’ve never done anything like that before. I guess I am a creative person, but I’m not an arty person, so it was a bit out of my comfort zone.’ (P01)*

Another (P05), found the task somewhat restrictive due to instructions limiting what content they could focus on, and one (P06) delayed the task, which seemed to stem from concerns about how others in the group may perceive their experiences:

> *‘I couldn’t decide what I wanted to talk about. At that time, I was aware of my online use and how negative it was when I was younger, there was still a stigma’ (P06)*

However, those who completed the task and presented it during Workshop 1, including members who were originally reluctant, named several benefits to involvement. This included giving and receiving positive feedback, and discovering new ways to express themselves. A few found the completion of erasure poems during Task 1 particularly helpful, as the constraints of the task made it easier to articulate complex thoughts and feelings:

> *‘Sometimes with these sensitive topics, if you’ve been through a lot there’s so much to say, that if you’re given text and you have to erase words and work with what you’ve got it forces you to express yourself a certain way… I was expecting it to be difficult because obviously you’re limited, but I thought it was a really good exercise.’ (P04)*

In Task 3, members were invited to write ‘search poems’ about their online experiences related to mental health. Several also enjoyed the reflective nature of this task:

> ‘it wasn’t creating as much as thinking, or forcing yourself to reflect on how you use the internet. It’s something you don’t really think about because we use it all the time, but I had to pick out certain things that were common threads for me… it forced me to reflect on the things that I’m actually searching for.’ (P04)However, one member (P05) had a more nuanced reaction. Whilst they recognised the beneficial nature of the reflection, this was balanced with the acknowledgement that revisiting these periods of poor mental health could have had a negative impact, if their personal resilience wasn’t as strong:
>
> *‘I think it would depend on someone’s mental state at the time. I can see how that might be slightly triggering, I mean it was quite sad for me to do. It also was a bit of a blur, the period I chose because I was quite unwell, but then it did help clarify that a bit.’ (P05)*
>
> Additionally, this task was described as ‘tricky’ (P06) due to the challenge of connecting online use with mental health, and P03 struggled with the directions. Whilst fewer members reflected on Tasks 2 and 4, P05 found the character development work (Task 4) was ‘really fun’, and P02 remarked ‘I really liked taking part in the survey (Task 2) too’ (P02).

#### Facilitators and Barriers to member involvement in the DDYPG

We identified several key factors that facilitated and caused barriers to successful involvement in the DDYPG.

##### Building Safety & Trust

Firstly, ensuring a safe and trusting environment was integral to DDYPG members’ involvement. Members expressed feeling: ‘it [was] a very safe space, safeguarding was great and was inclusive to all’ (anon1), and the requirement to complete a safety plan before involvement reassured members, ‘it was good to have that precaution*’* (P06). Others’ noted how completing a safety plan would be a *‘*good idea’ (P02, P03) for any mental health research involving young people, and one identified how it helped build rapport between the researcher and member:

> *‘It’s always good to have a safety plan for the young person but also the person doing the research because then at least you have that mutual understanding of what can be helpful and unhelpful during the involvement.’ (P03)*

None of the DDYPG members reported needing to access the safety plan during the study. This aligned with their self-perception of being comfortable and ‘confident’ (P06) discussing sensitive topics.

##### Positive Group Dynamics

The perception of safety was reinforced by the positive group dynamics. Members particularly appreciated the ‘non-judgemental’ attitudes from peers (P01, P03). The mutual awareness and understanding of handling potentially harmful information also played a role:

> *‘Luckily everyone else in the group was probably quite aware of sensitive topics we were discussing and perhaps not going into too much unnecessary detail that might be triggering. So I’ve never felt super uncomfortable.’ (P05)*

An additional factor that influenced DDYPG involvement was the opportunity for members to engage with peers without researchers present. This facilitated open conversations and allowed for organic idea generation:

> *‘I liked how we went into breakout rooms without the researchers, it felt like we were just talking young person to young person. Although the researchers are here and they understand the topic and they want to make a difference and make a change, a lot of the time they won’t have had these experiences before, sometimes that can make it difficult to talk to them.’ (P06)*

Members also emphasised the value of contributing to research that could help others. This sense of purpose encouraged their involvement and made them feel connected to a group of like-minded individuals:

> *‘it was good to talk to young people who want to be involved in a project to make a difference, and therefore are happy to talk about and share their experiences.’ (P06)*

##### Valued Members of the Project Team

DDYPG members consistently mentioned the quality of their involvement in the project as a significant motivator. The supportive relationships with researchers were a key factor, ‘I felt very cared for and valued.’ (anon2), and researcher responsiveness also played a role:

> *‘There were times where I would send you these huge rants in emails of all this stuff I noticed online, and you [the researcher] made sure I felt validated and I felt heard, which is really important for me.’ (P02)*

Additionally, the high level of DDYPG involvement in the study process was crucial. P01 remarked, ‘I feel like we’ve genuinely been quite equal partners in all of it, which is really cool’ (P01). This level of involvement was directly compared to other co-creation roles the group had been involved in: ‘ [The DDYPG] were different to other young people’s co-creation, they had a variety of different methods and options to choose from. I didn’t feel limited in any way.*’* (anon1). This promoted a sense of empowerment and encouraged the DDYPG members to question their other co-creation roles:

> *‘The level of involvement we’ve had has made me challenge a little bit [in other co-creation roles], like, ‘Why can’t we have more involvement? Why can’t we be doing this? Why can’t we be involved in that?’ (P01)*.

##### Time Management

Scheduling flexibility also facilitated involvement in this project and individual tasks. Members generally felt that their commitment to the DDYPG was *‘*manageable*’* (P01), as *‘*involvement was fairly spaced out’ (P03) and they could ‘balance’ (P04) it with other work including university and jobs. Additionally, the ability to continue conversations around task work on the Discord platform facilitated this flexibility:

> ‘*I appreciated the opportunity to participate in the tasks but then not necessarily have to be in the meetings to have discussions because they could move to Discord… I felt like we had good opportunities to participate in various different ways.’ (P01)*

A few members also noted that if tasks had shorter deadlines or were set all at once it may have been *‘*overwhelming’ (P03) and could have hindered their involvement. However, one member (P04) did feel they missed out on some involvement due to university obligations.

A couple of members observed that it was their responsibility to manage time and assess their capacity to complete DDYPG tasks alongside their daily lives. One recalled declining involvement comfortably:

> *‘There was a time I remember where you sent two tasks, and I was like to be completely honest I only really have time to do one, and you were like that’s absolutely fine just do the one. That was quite nice, as much as I wanted to do the other one I had to be realistic, you know, I’ve got a bunch of exams coming up, I don’t know if I can do both of those.’ (P02)*

However, the other felt less comfortable rejecting tasks, though they appreciated that presenting them as optional made decision-making less pressured:

> *‘you gave me the option, “would you like to do this” rather than “we’re going to do this”, I felt more able to say no. Although I never said no because I liked the project and I wanted to be involved, but I did like that I had the opportunity to say no or later on down the line I could be like “I don’t have time to do this, I’m sorry.”’ (P06)*

##### Anxiety

Most members expressed initial anxiety about attending the first DDYPG meeting, which could have acted as a barrier to involvement. Whilst two (P02, P03) attributed their apprehension to social anxiety diagnoses, others (P01, P03, P05, anon1, P06) shared similar concerns. They mentioned unfamiliarity with group members, fears that conversations might be triggering, and anxiety about presenting their creative work, especially when comparing it to the unknown pieces others had produced. However, all of these members also mentioned the anxiety quickly dissipating once the first meeting began:

> *‘I’d say just the nervousness of going on a zoom call with loads of people I don’t know and wondering if it’s going to be triggering or if it’s going to have an impact. And the nervousness of taking a piece of art and wondering what is this going to look like compared to everyone else…but I think that disappeared within five minutes of being on the call because everyone was just so nice and it was great to get to know everyone a little bit.’ (P01)*

Another concern that was mentioned by a couple of members, was that their experiences wouldn’t align with the group’s ‘norm’ regarding mental health and online use:

> *‘what if my idea about being chronically online and how harmful it is isn’t the norm?’ (P06)*

Additionally, one member expressed concern that their perspective might be ‘a really bad representation of people’s experiences’ (P05). This worry persisted throughout the project, as they explained:

> *‘It was in the back of my head that I didn’t want to say something—not wrong, but different or not representative enough’ (P05)*.

#### Member involvement in the DDYPG: Benefits and Risk

##### Validating Experiences

One of the primary benefits identified by DDYPG members was the opportunity to engage with individuals who had similar stories to them. This was seen as a chance to honestly talk about their mental health and online use (‘I felt positive about being able to share my experiences’ (anon2)) and hear from others, which many described as ‘validating’ (P01, P02, P04, P06). One communicated how this shared understanding helped them gain deeper insights into their own experiences:

> *‘I really enjoyed seeing the perspectives of people who’d been in similar situations to me and that helped me understand that side of using the internet in relation to my mental health a bit more.’ (P04)*

Young people also appreciated the chance to engage with peers who may have had different experiences to them, finding it valuable and ‘interesting’ (P04) to ‘[learn] more about other people’s perspectives’ (P01). This not only broadened their understanding of mental health, it helped them challenge their own preconceptions:

> *‘it was cool to know more things about them [DDYPG members’ mental health conditions], and probably addressing some of my own assumptions about them too…’ (P05)*

##### Positive Change to Online Behaviours and Mental Health

Additionally, involvement in the DDYPG led some members to reflect on and adjust their own behaviours to become more deliberate with how they navigated the online world, such as through spending less time online or changing the content they engaged with. For instance, one member stated:

> *‘I became more reflective about how I use my time online. I’m someone who likes to do scrolling like everyone else, so it felt a bit more intentional.’ (P02)*

Another noted that they started to critically evaluate other online users, which impacted their time spent online:

> *‘I noticed in my [online] use as well, that person is doing that that doesn’t make them a very good influencer, so thinking about this [research] was impacting my use too.’ (P06)*

Some of the members also reflected on the potential ‘therapeutic’ (P01) value of involvement in Digital Dialogues, specifically in the creative tasks. One shared how writing the ‘search poem’ allowed them to access and reconnect with their mental state during a difficult time, which had an overall positive impact:

> *‘My poem was about self-harming, and I think about it from my perspective now quite logically but my poem was that voice from when I was going through it. That made me connect to that situation more. I went back to how I was feeling rather than trying to intellectualise it… Just going back to how I felt and what it meant and why it happened, that was difficult but quite therapeutic and overall positive.’ (P04)*

Another member, who had some previous experience using creative methods to support their mental health, valued the option to explore a new outlet:

> *‘I’ve never really written poetry it was kind of therapeutic and I have now considered it.’ (P05)*

Another started using poetry as a therapeutic tool, as a direct result of their involvement:

> *‘I tend to write poetry now… Sometimes it’s around online use and sometimes generally mental health but I hadn’t thought about using creative outlets like poems until after I’d started in the Digital Dialogues project.’ (P06)*

##### Personal Development Opportunities

DDYPG members also highlighted how their involvement in Digital Dialogues positively impacted their individual development. P02 noted that being listened to and seeing their contributions used gave them self-assurance:

> *‘I think it helped my confidence quite a lot. Like I’ve said, knowing that my opinions were being heard and valued and they weren’t just thoughts I have that would fall on deaf ears and would never really make a change or anything.’ (P02)*

P03 also felt valued during the study, and pride in their role:

> *‘being involved has allowed me to feel like I’ve had a sense of purpose and more fulfilment in life. It’s helped with my general mood and feeling like I’m actually trying to make a difference. That’s the main thing that’s been positive, just that sort of feeling that I’m doing something that’s productive.’ (P03)*

Interestingly, three DDYPG members also mentioned how involvement in the project helped them overcome internalised stigma, which had previously stopped them talking openly about their mental health and online use.

> *‘It was difficult talking to people [in the group] originally about my experiences because I’d had this negative experience [talking to friends] in the past. But that was cleared up as soon as people started talking and I was like it’s not just a me thing, other people have experienced this and I’m not alone in this situation.’ (P08)*

##### Triggering Effect Of Conversations

Members identified that being in this project could also involve risks, including them being triggered by mental health related discussion. One, P02, shared that involvement in the study heightened their awareness of the online world, which left them more inclined to occasionally attend to potentially harmful content:

> *‘I guess on the negative side, particularly things about suicide these things are darker and deeper than it may appear to be, so when you notice that it can make you feel a bit sad.’ (P02)*

Additionally, P05 reflected on the potential negative impact that discussions around specific platforms or content could have on, noting that this may be dependent on their stage of recovery:

> *‘I would still say I’m recovering from an eating disorder so to be given a list of like “so I had difficulty with these specific forums or these websites”, if I was worse I probably would have looked them up. You have no way of knowing with all the other participants what level of recovery they’re at and if they might use that as a source… I definitely think it could have potentially done that for some people… I think there’s definitely a risk there.’ (P05)*

This member suggested researchers ‘ask people to explicitly avoid naming websites’ (P05) to avoid these triggers during conversations.

P01 also acknowledged that comparison to other members and triggering content were inherent risks in such discussions, but felt these were managed well in the project through the use of content warnings and the availability of researchers:

> *‘there were aspects of that that were a bit like oh okay this doesn’t feel quite so nice, and I think that’s always a potential when working with other people with that comparison and that triggering element. But I think overall, that was managed really well in terms of having trigger and content warnings and researchers in the meeting to talk to separately. So I don’t think it’s had any negative impacts on me.’ (P01)*

#### Young Person Reflections on Resource Development

##### Thoughts on the Resource Development Process

The resource development period was viewed positively by members, such as P02, ‘I think I most enjoyed creating the resources.’ (P02) and anon1, ‘it had a great positive impact, I felt included, heard and seen’ (anon1). Before beginning this part of the project, members were asked to identify the types of resources they would be interested in working on. Following this information, ZH approached members of the DDYPG to contribute either together or individually on the different stages of resource development. Members of the group who worked on specific tasks gave some reflections.

P05 described developing the video script alongside P01, highlighting how they were able to bring their own lived experience to the work and felt free to give honest input on the existing script. P05 appreciated the collaborative atmosphere, where they could engage critically whilst also sharing moments of humour related to their online use and mental health:

> *‘It was good to do the scriptwriting with [P01] too, that was really interesting. I enjoyed the conversation because it was funny and we could have a laugh, but also we were able to be quite critical of the script. Again, some of my ideas were probably quite different to her and that reflects how everyone’s experiences are very different.’ (P05)*

P01 also reflected on this collaborative relationship, noting the value of both being able to contribute their own perspectives:

> *‘The fact [P05] did the video script with me, I think it was really nice that we were the ones who had that kind of experience so we got to do the script-writing.’ (P01)*

P05 also described their involvement in the actor audition process, noting, ‘[I found] auditioning the actor really fun, I’ve never had to audition someone before and I really enjoyed that actually.’ (P05). This involvement also prompted a deeper reflection on representation of mental health in resources, such as those we created:

> *‘The last thing I wanted to do [whilst writing the script] was stereotype. I think that’s why it was important I was there for the auditions because I think some candidates erred on that side of it becoming a bit of a caricature, which we didn’t really want, and it also helped me think a bit more critically about portrayals of mental health.’ (P05)*

P06 also reflected on their individual role in designing and developing a poster and question bank directed at MHP’s having comfortable conversations with young people around their mental health and online use. They noted that this was not an easy process for them, due to concerns it would not be what the group hoped for:

> *‘The question bank as well. To me the question bank was really important, which is why it took me so long to do, I procrastinated on it for so long because I felt like it needed to be perfect.’ (P06)*

In addition, members generally reported their appreciation for the diverse roles they were able to have during resource development:

> *‘I think it was great to give young people choice and options to co-create through a variety of means and at a time and pace that works for them.’ (anon1)*

##### CYP Perception of Resource Use by MHPs

Five members expressed hopes that the resources created would provide an opportunity to improve the experience of CYP accessing support from MHPs. P03 reflected on the potential for MHPs to use them as communication aids, facilitating conversations:

> *‘I hope it’ll build communication and help MHPs to be a bit more comforting with the language that is used and the questions that are asked. I’m hoping it’ll be a good way to educate MHPs on how they can support younger people, as that’s the main aim of it, and hopefully the outcome.’ (P03)*

P05 expressed similar hopes, suggesting that resources could provide practitioners, specifically those working in CAMHS, with a ‘different lens’ through which they could understand and talk to young people about online use:

> *‘I’m hoping these will be a great prompt for people to take into their own practice and use to make young people feel more comfortable and not judged, because ultimately that’ll be the difference between them engaging with you and completely not.’ (P05)*

However, one member noted that the plans for disseminating resources to professionals were unclear to them, which meant they were uncertain about the potential impact:

> *‘I guess I wasn’t entirely sure what the plans were in terms of how you send them out. As in, is it every mental health professional, how is that possible? How do you even begin a task like that? That’s the only slightly grey area that once we’ve made these things, I wasn’t entirely sure how they would then get sent to people.’ (P02)*

## Discussion

Using creative methods, the Digital Dialogues project engaged young people in a research group where they shared their experiences and perspectives on online use and mental health. These discussions resulted in the iterative development of resources for practitioners, a virtual exhibition of creative works, and additional outputs. Findings from interviews with DDYPG members revealed key motivators for participation included creative engagement, quality of involvement, and peer interaction, which contributed to perceived benefits such as personal development, empowerment, and positive therapeutic outcomes. However, anxiety and time demands were identified as potential barriers to involvement, along with risks such as exposure to triggering or harmful content. Notably, the data also provide some evidence of steps that helped mitigate these barriers, allowing us to highlight key ethical considerations and potential strategies for future resource development projects.

### The Creative Process

Creative methods enabled young people to articulate their perspectives of online use and mental health within the group. This approach became an effective way to explore complex emotions and experiences [17]. Generally, CYP appreciated the novel approach, with high levels of task engagement. This was consistent with research showing that creative methods can enhance research involvement, by offering alternative forms of expression [19]. DDYPG members also described how they particularly valued constraint-based creative tasks, such as poetry writing. Here, structure helped them to describe their personal stories and communicate emotions that might otherwise remain intangible [20]. Additionally, young people reported that reflective aspects of creative tasks led to positive changes in online behaviours. This reflects findings from the DELVE study [20] where increased metacognitive skills led to positive behavioural changes online.

However, several DDYPG members also reported initial anxiety around producing or sharing their creative work, reflecting what Hochman and Esteves [22] term ‘art fear’. This also echoed observations by Novak-Leonard et al [23], that individuals with limited perceptions of themselves as an artist are less likely to engage in arts-based activities. To address this challenge, we adopted several strategies that had a positive impact on facilitating involvement and reducing negative emotions around the creative tasks. Firstly, we gave members flexibility by allowing them to use their preferred creative method to complete Task 1. This gave them the opportunity to draw on their strengths and work in a familiar way, using an assets-based approach [24].Similarly, tasks were framed as reflective rather than evaluative, aiming to mitigate performance-related anxiety [25]. Finally, we modelled involvement by having researchers share their own work first, a practice highlighted by Leavy [26] as effective in normalising creative engagement, and reducing power imbalances.

### Relationships Within the Research Team

A key facilitating factor for member involvement included the positive peer relationships they built and were able to rely on during the project. This led to feelings of safety, recognition, and validation amongst the young people, which may have enhanced engagement and confidence [27]. This was further reflected through members’ appreciation for opportunities to work independently with their peers. This movement away from researcher-led formats of collaboration better recognises the competency of CYP, and may also improve their commitment to the research [28]. However, although young people were able to work and contribute individually to resources, some hesitated to share their input due to concerns about not meeting group expectations or fear of being judged. This anxiety could sometimes delay contributions, and may have led members to withhold valuable ideas. Such challenges have been reported in previous research [29].

Rapport between DDYPG members and researchers was also integral to the young people’s active involvement in this project. Members reported that researchers were approachable and responsive, facilitating ongoing discussion, taking them seriously, and making them feel safe. This highlights the importance of researchers having the skills necessary to effectively collaborate with young people in their research, demonstrating a genuine commitment to authentic engagement, addressing power imbalances, and dedicating time to meaningful interactions [15].

### Ways of Working

Members also valued the responsibility and trust placed on them in their roles on this project, as well as the flexibility to contribute through various means. Other studies have also shown that offering several options for involvement in engagement work can improve inclusion [30], and using online communication platforms, such as Discord, can enhance this collaboration [16]. Additionally, members reported feelings of empowerment comparable to those experienced by others who have participated in meaningful co-production projects [31]. Further, our members highlighted flaws in other collaborative roles they had undertaken, revealing how effective collaboration can inspire critical reflection on past experiences, and empower CYP to challenge insufficient involvement.

Our working approach adhered closely to the principles outlined in the *Guidelines for Research with Children and Young People* [14]. Specifically, the development phase followed the ‘*CYP have ownership of the research’* model, which emphasised providing CYP with as much agency as possible. This approach empowered members, giving them a sense of fulfilment in their role. However, despite researchers’ efforts to maintain a manageable workload for CYP, one individual reported occasionally taking on more work than they could accommodate, driven by their enthusiasm for the study and desire to contribute. This emphasises the need for researchers to continually balance giving CYP agency with protecting their wellbeing [28].

### Managing Sensitive Content Discussions

DDYPG members appreciated how peers were mindful during discussions to avoid triggering content, interpreting this as a skilful use of boundaries grounded in a shared understanding of mental health challenges. Additionally, they recognised that their own stage of recovery was likely a key factor in their ability to cope with the discussions and tasks. This may reflect our group composition, as the recruitment strategy targeted CYP within mental health organisations or groups, where prior experiences may have helped them develop skills in navigating boundaries, addressing sensitive topics, and working collaboratively. It could also reflect the influence of group rules introduced and discussed during the initial workshop.

However, some members noted that despite efforts to avoid triggering content, information that could be potentially harmful was still shared. This suggests that the nature of conversations about mental health and online use may inherently involve exploring difficult or potentially triggering topics, which presents a challenge for researchers in balancing open dialogue and the emotional safety of CYP. Considering participants’ recovery stage during recruitment may therefore be an important factor. Research shows those with lived experience of mental health conditions experience varying levels of hope, meaning, confidence and symptoms at different stages of recovery, likely meaning they are able to contribute and cope to varying extents in research roles [32].

In an attempt to overcome potential risks of triggering content in this study, we provided opportunities for members to take breaks, access researchers for support during discussions in separate virtual breakout rooms, and ensured post-meeting check-ins. However, it remains unclear whether there may be longer-term negative or positive effects to CYP wellbeing or behaviours as a result of their involvement in research of this nature.

### Adhering to Perceived Norms

Some members expressed concerns about accurately representing their mental health experiences, reporting a perceived pressure to conform to a ‘norm’ associated with their diagnosis. Notably, this conformity to align with a mental health identity has recently been observed in individuals using social media, where online moderation and in-group formation play key roles in reinforcing diagnostic ‘norms’, particularly amongst young people [33–34]. These findings also reflect broader concerns with research engagement, such as the influence of Western societal expectations and desirability biases on participants’ willingness to engage in honest disclosures during mental health discussions [35]. Additionally, our efforts to minimise harm by introducing rules to avoid discussing triggering content may have created pressure for members to conform to a sanitised narrative.

Therefore, the inclusion of CYP with diverse mental health conditions had the potential to create a dynamic where individuals with less common diagnoses felt pressure to represent their condition. However, whilst this was an anticipated concern amongst members, it did not appear to be an influence once they took part in tasks and workshops. The group diversity also provided benefits, offering valuable peer learning opportunities and contributing to a potentially destigmatising environment. This supports research suggesting that diversity in groups can encourage broader perspectives and reduce stigma by exposing individuals to varied lived experiences [36]. Similarly, such diversity may enhance the generalisability of research insights by incorporating a wider range of perspectives.

### Perceptions of Project Outcomes

Members gained confidence from their involvement in this study, reflecting the concept that seeing ideas transformed into practical and tangible outcomes, is empowering [37]. They took pride in the created resources and felt hope that they would have an impact on MHPs, improving the ways they speak to CYP about their online use and mental health. However, members noted gaps in their understanding of how we planned to disseminate the resources to MHPs, a feature previously highlighted as important in collaborative research with young people [38].

### Limitations

This project successfully engaged young people as active contributors to the research through open discussions and creative work. DDYPG members played a key role in developing several resources for mental health practitioners. However, the following limitations highlight areas for reflection and potential improvements in resource development work with young people.

Members in this study generally reported a willingness to talk about their mental health with others, which contributed to the open and constructive discussions possible within the group. However, this also highlighted a potential self-selection bias in this type of research where those more comfortable discussing sensitive topics are more likely to be involved, and individuals who are less inclined to talk about their experiences may be underrepresented [15]. We tried to overcome this limitation through allowing CYP to be involved in the study in a variety of ways, including over an online discussion platform (Discord) and via commenting on and editing documents.

Additionally, one member noted uncertainty about the process for disseminating the resources to MHPs. Whilst this was a general limitation of the project, due to the need for additional funding to support this stage, it is important to consider that the lack of a clear dissemination plan from the outset may have reduced CYP’s sense of ownership or purpose in relation to the resources.

Finally, not all members participated in the evaluation interviews or the anonymous survey. Due to the anonymity of the survey, we cannot confirm whether those who completed it differed from those who took part in the interviews. As a result, we may have missed valuable perspectives from some members that could have provided additional insights.

## Conclusion

Involving young people with lived and living experience of mental health difficulties as research team members in a resource development project, can be mutually beneficial for both researchers and members. Using a structured format of workshops and creative tasks can encourage active involvement, and result in collaboratively conceptualised, designed, and created resources with an enhanced level of authenticity. According to DDYPG members, their role in this project was associated with positive outcomes, including empowerment, improved mental health, and a sense of validation. To enable this, it was important researchers created a safe space, and encouraged CYP agency and ownership over project decisions. However, challenges remained, including exposure to potentially triggering content, fear of judgement, anxiety around participation, and concerns about the impact of the developed resources. Through this evaluation, we have identified several mechanisms, as highlighted by CYP, to navigate and overcome some of these difficulties.

## Data Availability

Anonymised interview transcript and qualitative survey response data is deposited within the University of Bristol Research Data Repository. ‘Registered’ level data access will be applied, with data being available to researchers on request to Dr Zoe Haime at. Audio-recordings and transcriptions of the DDYPG workshops are closed and will not be available to share with other researchers, as agreed with the University of Bristol Data Protection services.

## Acknowledgements

We express gratitude to the DDYPG members for their contribution to this work. Appreciation also extends to networks that facilitated recruitment to the DDYPG. We would also like to thank our funders Research England Policy Support Fund. LB is partly funded by National Institute for Health Research Applied Research Collaboration West (NIHR ARC West) at University Hospitals Bristol NHS Foundation Trust. ZH is partly funded by Biomedical Research Centre, University Hospitals Bristol and Weston NHS Foundation Trust, Bristol, UK. HB is supported by an NIHR Advanced Fellowship (NIHR302271). The views expressed are those of the author(s) and not necessarily those of their affiliations. All authors have no competing or potential conflicts of interest.

